# Bio-Rad and QIAGEN digital PCR platforms provide equivalent quantification for wastewater-based SARS-CoV-2 surveillance

**DOI:** 10.64898/2026.01.20.26344437

**Authors:** Thomas Clerkin, Steph Smith, Kevin Zhu, Denene Blackwood, Javier Gallard-Góngora, Drew Capone, Joe Brown, Rachel T. Noble

## Abstract

Digital PCR (dPCR) is increasingly used for SARS-CoV-2 wastewater surveillance due to its precision, absolute quantification, and reduced sensitivity to inhibition compared to quantitative PCR. Although the Bio-Rad ddPCR and QIAGEN QIAcuity dPCR platforms are widely adopted, their performance has not been directly compared for wastewater applications. We conducted a blinded comparison of these platforms using 95 archived wastewater influent samples from North Carolina collected in 2021-2022, spanning three orders of magnitude in SARS-CoV-2 concentration (1×10^3^ to 5×10^5^ copies L^-1^). Samples were stratified into low, medium, and high concentration bins and analyzed in triplicate for N1 and N2 gene targets and a bovine coronavirus processing control. Both platforms demonstrated statistically equivalent quantification across all targets, with mean differences ≤0.12 log copies L^-1^ (R^2^ > 0.93). Coefficients of variation were similar (3.96 – 7.61%), with no significant differences across concentration bins except for N2 in the low bin (difference: 0.87 percentage points). Measurement variability correlated strongly with wastewater treatment plant site (R^2^ = 0.89) rather than platform, indicating that sample matrix characteristics drive precision more than analytical platform. Process limits of detection ranged from 2,160-2,680 copies L^-1^ for Bio-Rad and 5,650-9,700 copies L^-1^ for QIAcuity for N1 and N2, respectively. The Bio-Rad platform processed samples 32% faster (305 vs. 435 minutes per 96 wells), while QIAcuity offered 29% lower consumables cost ($4.68 vs. $6.11 per well). These findings support the interchangeable use of both platforms for wastewater surveillance, with platform selection based on laboratory-specific operational needs.

**Importance:** As wastewater-based epidemiology transitions from emergency response to sustained public health infrastructure, standardized molecular methods are essential for reliable data integration across surveillance networks. This study provides the first blinded comparison of two digital PCR platforms widely deployed for wastewater pathogen surveillance in the United States. We demonstrate quantitative equivalence between Bio-Rad ddPCR and QIAGEN QIAcuity platforms across three orders of magnitude in viral concentration, establishing that data from both platforms can be interpreted interchangeably for public health decision-making. This platform equivalence is critical as national surveillance systems aggregate data from diverse laboratories and as monitoring expands beyond SARS-CoV-2 to encompass additional respiratory viruses, antimicrobial resistance genes, and emerging pathogens. Our findings provide a methodological foundation for multi-platform surveillance networks and demonstrate that measurement variability is driven primarily by sample matrix characteristics rather than analytical platform choice.

## Introduction

Wastewater-based epidemiology emerged as a critical public health tool during the COVID-19 pandemic, with over 4,000 monitoring sites now operating across more than 70 countries worldwide (1). Although wastewater surveillance was developed in the 20^th^ century, the pandemic accelerated its global adoption through rapid mobilization of the academic and public health research communities (2). In the United States, this expansion has been formalized through the establishment of the Centers for Disease Control and Prevention (CDC) National Wastewater Surveillance System (NWSS), which integrates wastewater data into routine public health monitoring and decision-making (3,4). As wastewater surveillance programs mature and expand beyond SARS-CoV-2 to encompass additional respiratory viruses, antimicrobial resistance genes, and emerging pathogens (5–10), standardizing analytical methods and ensuring data comparability across platforms and laboratories has become increasingly important for reliable public health decision-making (3,10–13).

Most wastewater surveillance programs quantify pathogen-specific nucleic acid sequences using PCR-based approaches. Early in the COVID-19 pandemic, both quantitative PCR (qPCR) and digital PCR (dPCR) were widely used, and comparative studies demonstrated that both methods could be effective for detecting SARS-CoV-2 in wastewater (14–18). However, since late 2022, dPCR has emerged as the preferred platform for wastewater surveillance due to several advantages that address challenges inherent to wastewater matrices. First dPCR partitions reactions into thousands of discrete volumes, enabling absolute quantification through Poisson statistics and providing superior precision at low target concentrations, particularly in matrices containing PCR inhibitors such as humic substances prevalent in wastewater (19,20). Second, dPCR eliminates the need for standard curves required by qPCR, reducing quality control requirements and simplifying workflows for high-throughput surveillance programs. Third, dPCR demonstrates greater tolerance to inhibition, minimizing sample dilution and reanalysis while maintaining sensitivity (16). Multiple comparative studies have demonstrated dPCR’s advantages over qPCR for wastewater applications, showing improved limits of detection, reduced variability, and more reliable quantification across diverse sample matrices (14,16,21,22). Together, these features have supported more consistent laboratory turnaround times and standardized workflows, facilitating near-real-time epidemiological interpretation, trend analysis, and public health response within surveillance systems such as the CDC NWSS (3,5,10–13,23).

Two dPCR platforms currently dominate wastewater surveillance of SARS-CoV-2 in the United States: the Bio-Rad QX200 and QX600 droplet digital PCR (ddPCR) systems and the QIAGEN QIAcuity digital PCR platforms. The Bio-Rad QX200, released in 2013, uses microfluidic technology to partition PCR reactions into approximately 20,000 nanoliter-scale droplets suspended in oil, which are individually analyzed for fluorescence after thermal cycling. The QIAGEN QIAcuity platform, introduced in 2020, employs pre-formed partitions in disposable nanoplates containing up to 26,000 partitions per well, fluorescence detected via imaging. Both platforms have been independently validated for SARS-CoV-2 wastewater surveillance and are supported by the CDC NWSS. Between 2021 and 2022, at least 50 publications were written regarding the implementation of dPCR for related applications of wastewater surveillance.

Numerous studies have demonstrated the utility of each platform individually for wastewater surveillance applications, and several investigations have compared qPCR-based and dPCR-based quantification of SARS-CoV-2 targets in wastewater (14–16,18). Soon after the implementation of widespread wastewater surveillance using dPCR, many additional laboratories, including contract, public, and private entities generated assays for and utilized dPCR as their preferred technology. However, despite their concurrent and widespread use, no studies to date have conducted a blinded, head-to-head comparison of these two digital PCR platforms for wastewater surveillance following established digital MIQE guidelines (24).

As wastewater surveillance transitions from an emergency response tool to a sustained component of public health infrastructure, confidence in analytical comparability across platforms is critical. National surveillance programs aggregate data generated across diverse workflows, instruments, and reagents, raising questions about the extent to which methodological differences may influence reported concentrations, trend interpretation, and downstream public health decision-making. Direct evaluation of platform-to-platform performance is therefore essential not only for methodological validation, but also for ensuring that surveillance data remain interpretable and actionable as these systems continue to expand.

The objective of this study was to conduct a blinded, systematic comparison of the Bio-Rad QX200 ddPCR and QIAGEN QIAcuity dPCR platforms for wastewater-based SARS-CoV-2 surveillance. Specifically, we evaluated: (1) quantification equivalence for SARS-CoV-2 N1 and N2 gene targets and bovine coronavirus processing controls across low, medium, and high concentration ranges using both single-well and hyperwelled (merged triplicate) approaches; (2) precision and reproducibility as assessed by coefficients of variation; (3) process limits of detection and quantification; (4) sensitivity to matrix-related inhibition effects; (5) the impact of fluorescence threshold positioning on quantification; and (6) practical considerations including workflow efficiency, processing time, and consumable costs. To accomplish these objectives, we analyzed 95 archived wastewater influent samples collected from multiple wastewater treatment plants across North Carolina during 2021-2022 as part of routine CDC NWSS monitoring. Samples were selected by a blinded researcher across a wide concentration range, extracted using standardized methods, and analyzed independently by two laboratories, each experts in their respective platform. While this study focuses on SARS-CoV-2 quantification, the rigorous comparison framework has broad relevance to the expanding applications of dPCR in wastewater surveillance, including detection of antimicrobial resistance genes (6,7), variant tracking (8), and emerging pathogens (9,10).

## Materials and Methods

### Sample Collection and Processing

Raw influent wastewater samples were collected as part of routine monitoring by the North Carolina National Wastewater Sewage Surveillance System (NC NWSS) with support from the North Carolina Department of Health and Human Services and United States Centers for Disease Control and Prevention. Wastewater treatment plant (WWTP) staff collected 250mL samples of 24-hour flow weighted raw influent wastewater in 250mL polypropylene sample bottles and stored them at 4°C. Samples were shipped overnight on refrigerant gel to the UNC Institute of Marine Sciences (IMS, Morehead City, NC) within 6 days of collection.

Immediately upon arrival, samples were adjusted to a pH of 3.5 as confirmed by pH test paper by adding up to 2mL of 10M hydrochloric acid (HCl) to the 250mL sample. Additionally, research analysts amended each sample by the addition of 5mL of 1.25 M magnesium chloride hexahydrate (MgCl_2_) to achieve a final concentration of 25mM. The acidification and amendment of samples with MgCl_2_ were performed to aid in viral adhesion to Mixed Cellulose Ester (MCE) membrane filters (25). Furthermore, each sample was spiked with 100,200 copies of Bovine Coronavirus (BCoV: MERECK Animal Health BOVILIS® Coronavirus Calf Vaccine, PBS Animal Health Massillon, OH) to act as a total processing control. Immediately following amendment, samples were mixed vigorously by inversion for 30 seconds and then 40mL of sample was vacuum filtered to dryness in disposable filter funnels through type GN-6 Metricel 47mm 0.45 μm MCE filters (Pall Corp., Port Washington, NY) in quadruplicate. Filters were carefully placed in 2mL microcentrifuge tubes and archived at-80°C for up to 4 months.

### Sample Selection and Blinding

From over 4,000 wastewater samples processed by the Noble Laboratory for CDC NWSS between March 2020 and February 2023, a research analyst not associated with this study used inventory records and previously reported N1 and N2 concentrations to randomly select 96 archived filters. Samples were stratified into three concentration bins based on arithmetic mean SARS-CoV-2 concentrations from routine ddPCR surveillance: low (n=31), medium (n=32), and high (n=31), plus two field blanks. Selected filters were assigned unique identification numbers using a random number generator (Microsoft Excel v.2302) to blind the analytical laboratories to sample identity and prior concentration data. After dPCR analyses were completed, sample identifiers were unblinded to permit statistical comparisons based on concentration bins, WWTP characteristics, and other sample metadata.

### Total Nucleic Acid Extraction and Reverse Transcription

Archived filters were thawed at room temperature for 10 minutes and submerged in 1 mL of EasyMag® Lysis buffer (bioMerieux, Durham, NC) containing 9,000 copies of Armored RNA® containing a portion of the human hepatitis G sequence (HepG, Asuragen Austin, TX) to assess the extraction recovery efficiency of each sample. Lysed samples were incubated at room temperature for 10 minutes and the supernatant was transferred to a 96 deep-well plate (95040450, Thermo Fisher, Waltham, MA) for automated total nucleic acid extraction on the KingFisher™ Flex (Thermo Fisher Scientific) with EasyMag® NucliSENS ® reagents (bioMerieux) as described by Beattie, et al., 2022. Only a single filter for each sample was processed. Detailed extraction protocols, reagent volumes, and KingFisher protocol parameters are provided in Supplemental Tables S1-S3 and Figure S1. Total nucleic acids were eluted in 100μL of Buffer AE (19077, QIAGEN, Germantown, MD) and transferred into 96-Well ddPCR Plates (12001925, Bio-Rad Laboratories).

A two-step reverse transcription (RT) protocol converted RNA to cDNA using the Reliance Select cDNA Synthesis kit (BioRad Laboratories). The RT mastermix contained 37.5μL of extracted RNA, 15μL of Reliance Select cDNA Synthesis Reaction Buffer, 3.75μL of Reliance Reverse Transcriptase, 7.5μL of 10X Reliance Random Primer Mix (BioRad Laboratories), 10.75 μL of DEPC-treated water (Thermo Fisher Scientific, Waltham, MA), and 0.5μL containing 2,000 copies/μL of Total RNA- Mouse l Lung Normal Tissue (mouse lung; R1334152-50, BioChain Institute, Newark, CA) to assess the RT efficiency of each sample, resulting in a total reaction volume of 75μL. A C1000 Touch™ thermal cycler (Bio-Rad Laboratories) was used to conduct the RT thermal cycling conditions according to the manufacturer’s instructions and as described in the supplementary information. After cDNA synthesis, duplicate reactions were combined and briefly vortexed for a total cDNA volume of 150μL per sample. Samples were then transferred into duplicate 0.5mL microcentrifuge tubes and stored at-80°C for up to 4 months. The Noble laboratory (IMS) kept one aliquot of cDNA for PCR analysis on the QX200 platform (Bio-Rad Laboratories) and the other aliquot was shipped on dry ice to the Brown Lab (UNC-CH, Chapel Hill, NC) for analysis on the QIAcuity Four (hereafter referred to as QIAcuity) digital platform (QIAGEN, Germantown, MD).

### Bio-Rad QX200 Droplet Digital PCR

CDC-recommended primer and probe sequences targeting SARS CoV-2 nucleocapsid N1 and N2 regions (33) were used in duplexed reactions (Table S5). Duplexed assays included: (1) N1 (FAM) with HepG (HEX) to assess extraction efficiency; (2) N2 (FAM) with Mouse Lung Beta Actin (VIC) to assess RT efficiency; and (3) BCoV (FAM) with *gyrA* gene from haloalkaliphilic archaeon (HEX) to assess total processing efficiency and PCR inhibition (Table S6). Complete primer and probe sequences are provided in Table S5.

PCR reactions contained 5µL cDNA template, 12.5µL of ddPCR^TM^ 2x Supermix for Probes (no dUTP, Bio-Rad), 0.9µM forward and reverse primers, 0.25µM probes, and nuclease-free water to 25µL final volume. For the BCoV/*gyrA* duplex, 1µL containing 60 copies of *gyrA* was added to assess PCR inhibition. All samples were run in triplicate. Detailed mastermix compositions are provided in Table S9. PCR mastermixes were loaded into an Automated Droplet Generator (AutoDG, Bio-Rad) to partition 20µL into approximately 20,000 droplets per well. Plates were sealed with pierceable foil (PX1^TM^ PCR Plate Sealer, Bio-Rad) and thermally cycled using conditions in Table S7.

### QIAGEN QIAcuity Four Digital PCR

Identical primer and probe sequences were used on the QIAcuity platform but at optimized concentrations determined through titration experiments (Table S8). Optimization aimed to achieve positive partition fluorescence of 75-150 relative fluorescence units (RFUs) and negative partition fluorescence <50 RFUs while maintaining consistent thermal cycling conditions (40 cycles, 55°C annealing) with the Bio-Rad platform. Final reaction concentrations varied by assay (Table S8).

PCR reactions contained 5µL of cDNA template, 10µL of 4x QIAcuity Probe Mastermix (QIAcuity Probe PCR Kit, QIAGEN), optimized concentrations of primers and probes, and UV-treated molecular-grade water to 40µL final volume. For the BCoV/*gyrA* duplex, 1µL containing 60 copies µL^-1^ of *gyrA* was added. All samples were run in triplicate. Detailed mastermix compositions are provided in Table S10. Master mixes (39µL) were loaded onto QIAcuity Nanoplate 26k 24-well plates (QIAGEN), sealed according to manufacturer instructions, and loaded into the QIAcuity Four. Thermal cycling conditions are reported in Table S8. Imaging was performed using Green (FAM) and Yellow (HEX/VIC) channels with 500 ms exposure and gain of 6.

### Digital PCR Fluorescence Amplitude Thresholding

Fluorescence amplitude thresholds were set identically for both platforms. Thresholds were positioned manually at the midpoint between the average fluorescence of positive and negative clusters identified in positive and negative controls, respectively. For the N1 assay, which exhibits multiple positive droplet populations due to viral mutations (26), the threshold was set at the midpoint between the average fluorescence of the lowest positive population and the negative population. This approach was applied consistently across all samples and both platforms. Representative threshold partitioning screenshots are shown in Figure S3.

## Data Analysis and Normalization

### Bio-Rad QX200

Thermal cycled droplets were read on a QX200 reader (Bio-Rad Laboratories) and analyzed using QuantaSoft™ v. 1.7 software (Bio-Rad Laboratories, Hercules, CA) which applies Poisson distribution statistics to quantify target concentrations in a sample based on positive, negative, and accepted droplets distributions. Wells with <10,000 accepted droplets were excluded per the manufacturer’s recommendations. One of the 96 samples analyzed was excluded due to insufficient droplet formation. The droplet data was exported to a spreadsheet using a CSV file and then used to calculate the concentration of the samples in copies per microliter in the ddPCR reaction, with an assumed droplet volume (0.85 nL). The concentration data was saved as a Microsoft Excel v.2302 workbook and normalized to copies 1L^-1^ influent wastewater using the formula described by Beattie, et al., 2022 (25) to account for the volume of sample filtered, elution volume, and the RT dilution factor. For the Bio-Rad QX200, samples were considered positive and included in statistical analyses if they were above the Analytical LOD of containing at least three positive droplets following the merging of replicate wells.

### QIAGEN QIAcuity Four

Thermally cycled QIAGEN 26k Nanoplates were imaged on the QIAcuity Four (QIAGEN) running the QIAcuity Software Suite 2.1.7.182 with QIAcuity Volume Precision Factor version 4.0. Imaging of FAM and HEX/VIC fluorophores was done using the Green and Yellow channels (respectively), with an exposure duration of 500 ms for all assays used in this study and gain of 6. dPCR wells were rerun if the reaction contained less than 16,000 accepted partitions (occurred in 4 instances). The concentration data was then exported as a CSV file and then used to calculate the concentration of the samples in copies per microliter in the dPCR reaction, with an approximate droplet volume (0.8nL). The concentration data was then normalized to copies L ^-1^ influent wastewater using the formula described by Beattie, et al., 2022 (25) to account for the volume of sample filtered, elution volume, and the RT dilution factor. For the QIAGEN QIAcuity, samples were considered positive if there were at least three positive partitions following the merging of replicate wells.

### Limit of Blank and Process Limit of Detection

The Limit of Blank (LOB) was determined using eight technical replicates of eight wastewater influent samples collected in 2018 before the COVID-19 pandemic that were negative for SARS-CoV-2. There was no positive fluorescence signal in either the N1 or N2 assays for both platforms, thus the LOB for each assay on both Bio-Rad and QIAGEN platforms was determined to be 0. For the analyzed wastewater samples, an Analytical Limit of Detection of three positive partitions per dPCR/ddPCR well was used to determine if a sample was positive or negative for an assay. To understand the Process Limit of Detection, or at what concentration normalized to liters of wastewater would result in a detection 95% of the time, a Probit Analysis adapted from the clinical guidance document entitled “Protocols for Determination of Limits of Detection and Limits of Quantitation; Approved Guideline” (Clinical and Laboratory Standards Institute document EP17-A2) (27). The guideline was followed to determine the Process Limit of Detection (PLOD) for a single dPCR or ddPCR well for N1 and N2 by running 30 replicates of six serial dilutions. Microsoft Excel v.2302 was used to determine the LOD with 95% confidence of 2160 copies L^-1^ and 2680 copies L^-1^ for N1 and N2, respectively on the Bio-Rad QX-200 platform and 5650 copies L^-1^ and 9700 copies L^-1^ for N1 and N2, respectively on the QIAGEN QIAcuity platform.

### Quality Control Elements

This study followed MIQE guidelines for digital PCR applications as specified in Huggett et al. 2020; see dMIQE checklist in Supplemental Table S14 (24). Supplemental Table S1 refers to all required quality control elements for both the QX200 and the QIAcuity Platforms. Here, we highlight specific elements of quality control of interest for manuscript interpretation, but many of the details for the quality control elements can be found in the SOI section.

Positive/Negative Controls: Bovine Coronavirus (BCoV: MERECK Animal Health BOVILIS® Coronavirus Calf Vaccine, PBS Animal Health Massillon, OH) was used as a positive processing control for this study. Total processing recovery efficiencies were determined via the method outlined in Ciesielski et al. 2020. The total processing efficiencies were assessed by spiking 100,200 copies of BCoV into each 250mL sample before filtration to quantify the percent recovery of viral RNA throughout the entire process. Total processing recovery derived from BCoV concentrations were calculated using the following equation:

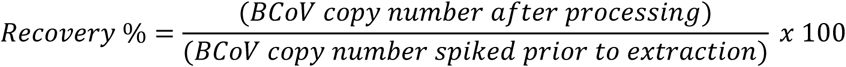

Calculated recoveries fell within a range of 0.56% and 50% and 0.17% and 29% for the Bio-Rad QX200 and the QIAGEN QIAcuity, respectively.

Extraction efficiency was calculated by measuring the percentage of Hepatitis G recovered after the extraction process as compared to the known amount added to the reaction. Inhibition during reverse transcription was assessed by comparing the known concentration of Mouse Lung RNA in the samples to the concentration found in the Negative Extraction Control (NEC). Additionally, inhibition during PCR was assessed by comparing the known concentration of the *gyrA* gene from the spiked in haloalkaliphilic archaeon and to the No Template Control (NTC). Samples were considered inhibited if the concentration of RT or PCR control was greater than 2 standard deviations away from the NEC or NTC, respectively. No samples in this study were determined to be inhibited.

This study verified the absence of contamination at each step of the process by including different types of negative controls. Two field blank samples (FB), four NECs, and at least four NTCs were included with each duplexed assay plate. Each FB consisted of a 250mL polypropylene bottle that was uncapped at a WWTP, filled with 250mL of water, placed at 4°C, and shipped and processed with other wastewater samples, including amendment and membrane filtration. Each FB was analyzed in triplicate with each assay. The NECs consisted of new MCE filters placed in 2mL microcentrifuge tubes and extracted along with the wastewater samples. The NECs were run in duplicate with each assay. A minimum of four NTCs composed of 5μL of nuclease-free water, containing no target analyte, were added to PCR mastermixes in place of cDNA template and run on each assay plate. All negative controls were negative during this study and there were no instances of contamination. For positive controls, we used genomic SARS-CoV-2 RNA positive control from strain 2019nCoV/USA-WA1/2020 (VR-1986D, ATCC Bethesda, MD), and the positive controls were included with each plate in duplicate for the N1 and N2 assays.

### Time and Cost Analysis

Workflow time was assessed by measuring hands-on time and instrument time required for skilled laboratory technicians (≥3 years platform-specific experience) to process 96 wells from mastermix preparation through data export. Times excluded upstream steps (collection, filtration, extraction, RT), which were identical between platforms. Consumable costs were calculated using manufacturer list prices (accessed December 4, 2024) for all items required to process 96 wells, excluding costs for sample processing, extraction, and RT reagents, which were platform-independent. Capital equipment, maintenance, and service contract costs were not included.

## Statistical Analysis

Concentration data (copies L^-1^) were log_10_-transformed to reduce skewness and approximate normality. Preliminary one-way ANOVA models assessed normality through residual diagnostics (histograms, Q-Q plots, Shapiro-Wilk tests). Due to deviations from normality, Aligned Rank Transform (ART) ANOVA was applied using the ARTool package in R. Separate ART models were fitted for each concentration bin (low, medium, high, all) with the form: value (aligned ranks) ∼ N x PCR, where N represents target (N1 vs. N2) and PCR represents platform (Bio-Rad vs. QIAcuity).

Linear mixed effects models evaluated platform differences in log_10_ concentrations using the lme4 package. Platform and concentration bin were entered as fixed effects, with sample identifier as a random effect. Tukey’s post-hoc tests compared differences between bins and platforms. Coefficient of variation (CoV) was calculated as (standard deviation / mean) x 100 for triplicate reactions. CoV equality between platforms was assessed using modified signed-likelihood ratio tests (cvequality package v.0.2.0). Statistical significance was determined using α = 0.05. Linear regression assessed platform correlations. All analyses were performed in R v.4.4.1 using packages: tidyverse, ggplot2, car, ARTool, lme4, and cvequality.

## Results

### Overall Platform Comparison

We evaluated 96 wastewater influent samples across concentration bins spanning three orders of magnitude (1×10^3^ to 5×10^5^ copies L^-1^) using both the Bio-Rad QX200 ddPCR and QIAGEN QIAcuity Four dPCR platforms. All samples were analyzed in triplicate for SARS-CoV-2 N1 and N2 gene targets and the bovine coronavirus (BCoV) processing control. One sample failed to generate sufficient droplets on the Bio-Rad platform (<10,000 accepted droplets) and was excluded from analysis, resulting in 95 samples analyzed on both platforms.

The two platforms yielded statistically equivalent quantification across all targets when the complete dataset was analyzed together (Table 2, Figure 1). The mean different in log_10_-transformed concentrations between platforms was 0.10 log copies L^-1^ for N1, 0.06 log copies L^-1^ for N2, and 0.12 log copies L^-1^ for BCoV. Linear regression analysis demonstrated strong correlation between platforms for all targets (R^2^ > 0.95 for N1 and N2, R^2^ > 0.93 for BCoV), with regression slopes near unity (0.97 – 1.02) indicating minimal systematic bias (Figure 1).

**Figure 1.**
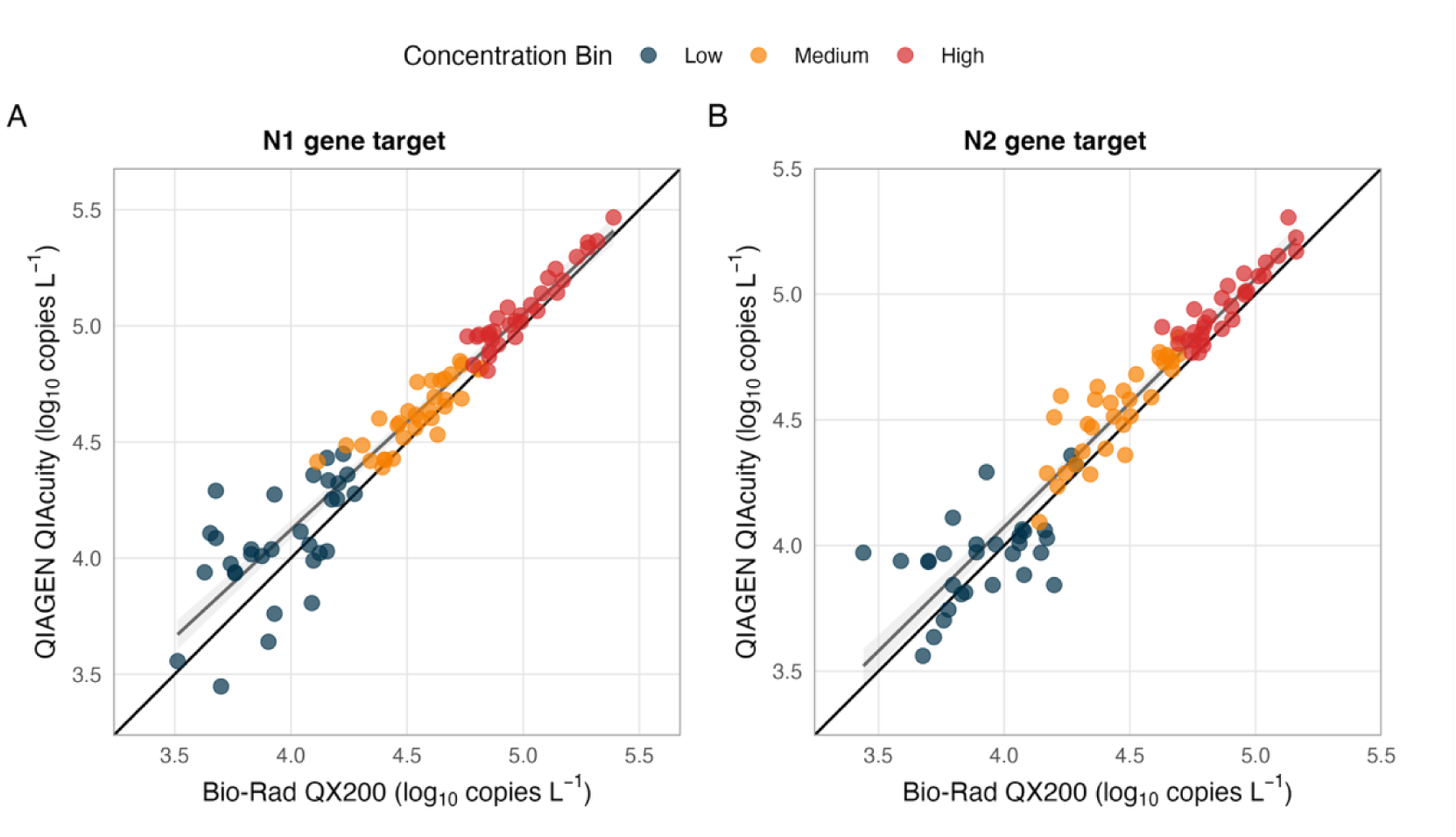
Quantitative comparison of SARS-CoV-2 targets between Bio-Rad QX200 ddPCR and QIAGEN QIAcuity dPCR platforms across all concentration ranges. Scatter plots showing hyperwelled (merged triplicate) concentrations for (A) N1 gene target and (B) N2 gene target. Each point represents a single wastewater sample (n = 95) plotted as log_10_ copies L^-1^ on both axes. Samples are color-coded by concentration bin: high (red, 5×10^4^ to 5×10^5^ copies L^-1^, n=31), medium (orange, 5×10^3^ to 5×10^4^ copies L^-1^, n=32), and low (blue, 1×10^3^ to 5×10^3^ copies L^-1^, n=31). Solid black line represents perfect 1:1 agreement; dashed line showed linear regression fit. Linear regression equations, R^2^ values, and 95% confidence intervals (gray shading) are displayed for each target. Mean absolute differences between platforms: N1 = 0.10 log copies L^-1^, N2 = 0.06 log copies L^-1^. All correlations significant at p < 0.001.

Aligned Rank Transform (ART) ANOVA revealed no significant interaction between platform and target gene (N1 vs. N2) across the entire dataset (p > 0.05), indicating that the platforms performed consistently regardless of which target was quantified. While rank distributions showed a slight but statistically significant difference between N1 and N2 quantification (p < 0.01), this difference was independent of platform choice and likely reflects biological variation in target copy numbers within the viral genome.

### Performance Across Concentration Ranges

To evaluate platform performance at different viral concentrations, we stratified samples into tertile bins based on arithmetic mean SARS-CoV-2 concentrations previously determined during routine surveillance: high (5×10^4^ to 5×10^5^ copies L^-1^, n=31), medium (5×10^3^ to 5×10^4^ copies L^-1^, n=32), and low (1×10^3^ to 5×10^3^ copies L^-1^, n=31) (Table 1). Both platforms successfully amplified and quantified all samples across all concentration bins with 100% detection rates.

**Table 1.**
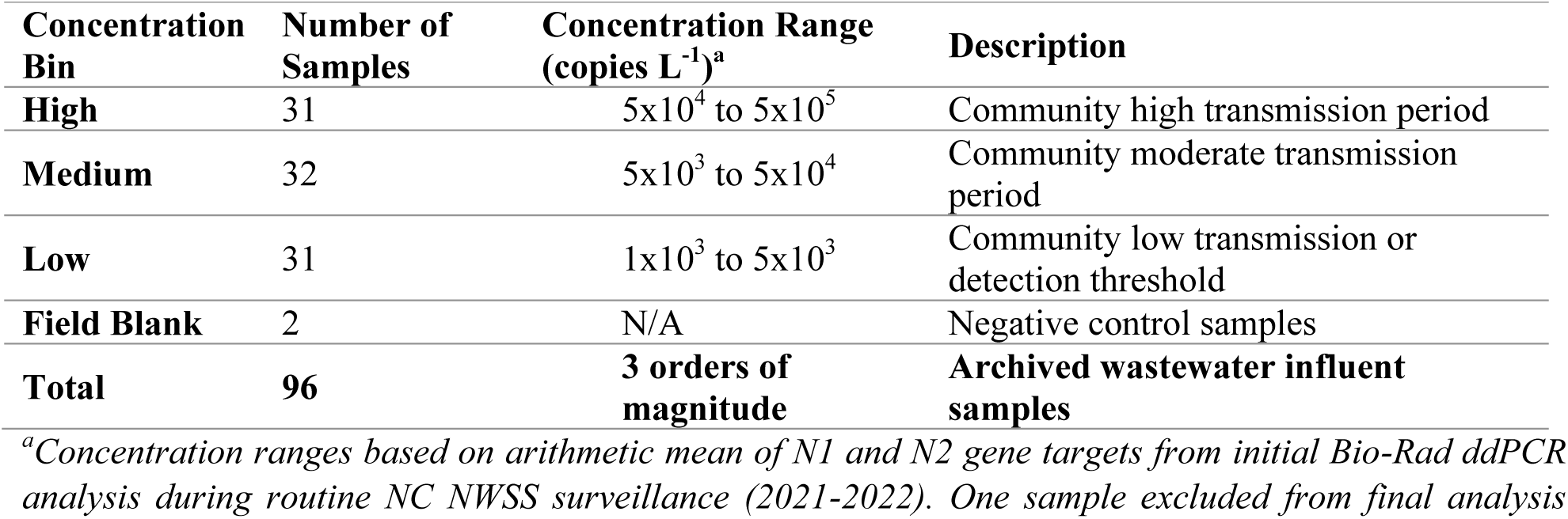
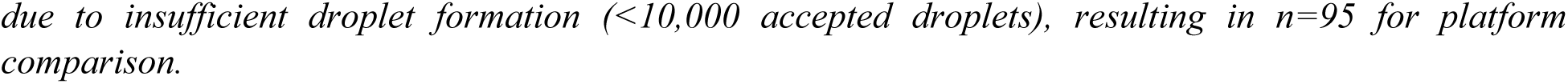
Sample distribution and concentration ranges used to platform comparison.

| Concentration Bin | Number of Samples | Concentration Range (copies $L^{-1}$)<sup>a</sup> | Description |
| --- | --- | --- | --- |
| High | 31 | $5 \times 10^4$ to $5 \times 10^5$ | Community high transmission period |
| Medium | 32 | $5 \times 10^3$ to $5 \times 10^4$ | Community moderate transmission period |
| Low | 31 | $1 \times 10^3$ to $5 \times 10^3$ | Community low transmission or detection threshold |
| Field Blank | 2 | N/A | Negative control samples |
| Total | 96 | 3 orders of magnitude | Archived wastewater influent samples |

<sup>a</sup>Concentration ranges based on arithmetic mean of N1 and N2 gene targets from initial Bio-Rad ddPCR analysis during routine NC NWSS surveillance (2021-2022). One sample excluded from final analysis
due to insufficient droplet formation (<10,000 accepted droplets), resulting in $n=95$ for platform comparison.

**Table 2.**
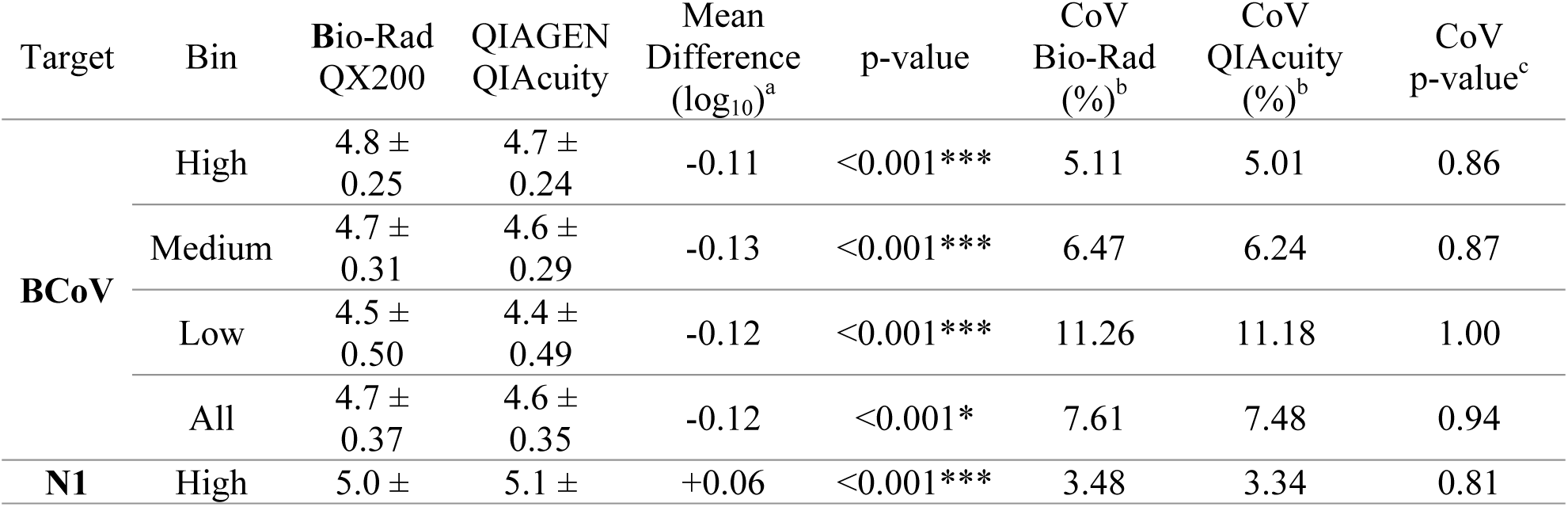
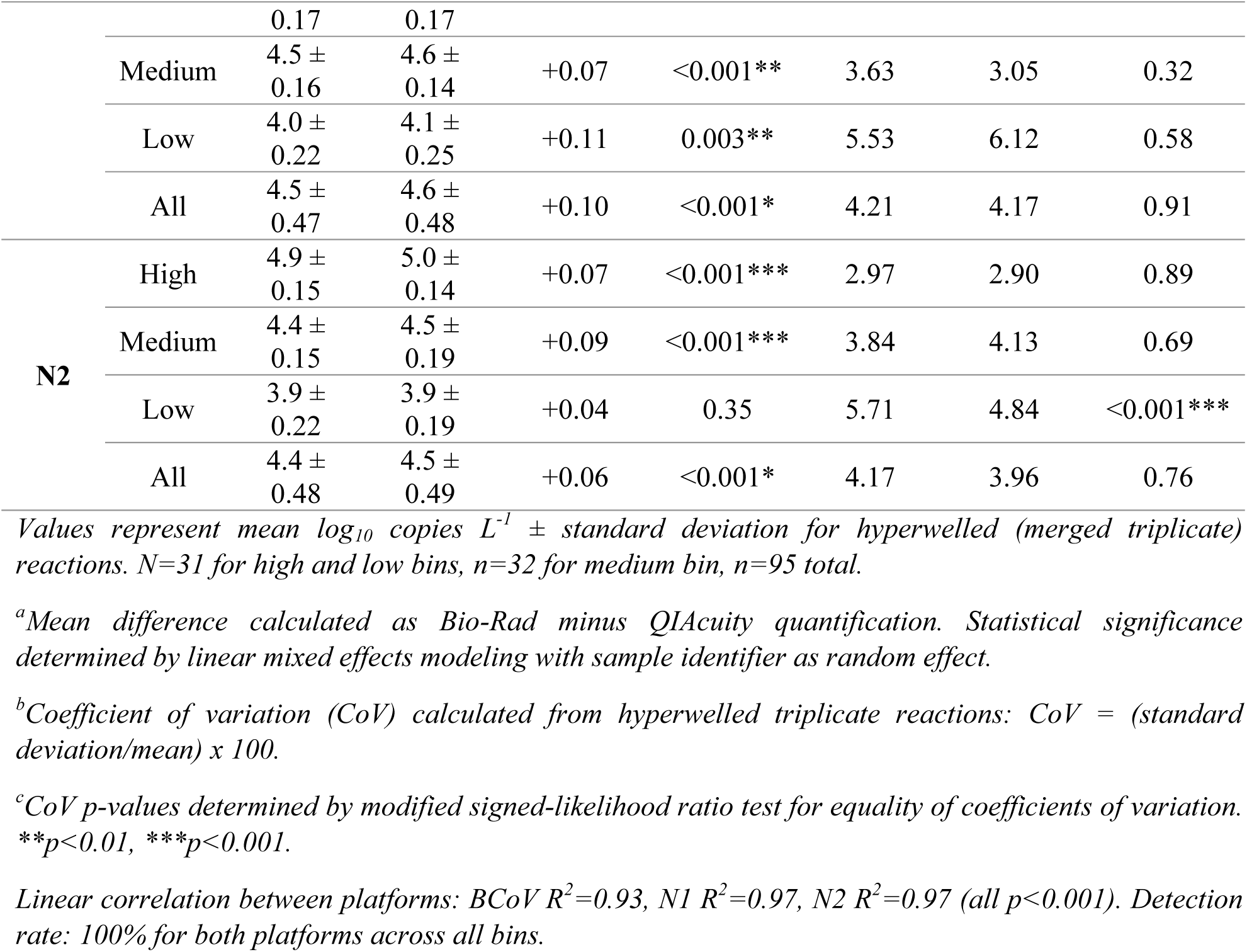
Quantitative comparison and precision metrics for Bio-Rad QX200 and QIAGEN QIAcuity platforms across concentration bins.

Linear mixed effects modeling revealed small but statistically significant differences in quantification between platforms when analyzed by concentration bin (Table 2). For N1, the Bio-Rad platform reported marginally higher concentrations across all bins (difference: 0.06-0.11 log copies L^-1^, p < 0.01), while for N2 this trend was observed only in high and medium bins (difference: 0.07 – 0.09 log copies L^-1^, p < 0.001) but not in the low concentration bin (difference: 0.04 log copies L^-1^, p = 0.35). The BCoV processing control showed the opposite pattern, with the QIAcuity platform reporting slightly higher concentrations across all bins (difference: 0.11 – 0.13 log copies L^-1^, p < 0.001).

Despite statistical significance, the magnitude of these differences (0.04 – 0.13 log copies L^-1^) represents less than 35% variation in absolute concentration values and falls well within the natural variability observed in wastewater surveillance programs. ART ANOVA by concentration bin confirmed that while differences existed between N1 and N2 quantification within bins, there was no significant interaction between platform and target (p > 0.05 for all bins), supporting platform equivalence for practical surveillance applications.

For the low concentration bin, neither platform showed significant differences in rank distributions (p > 0.05), indicating robust performance even near the limits of detection. In medium and high concentration bins, while platforms showed statistically different rank distributions (p < 0.05), the effect sizes were small and neither platform consistently outperformed the other across all targets (Figure 2).

**Figure 2.**
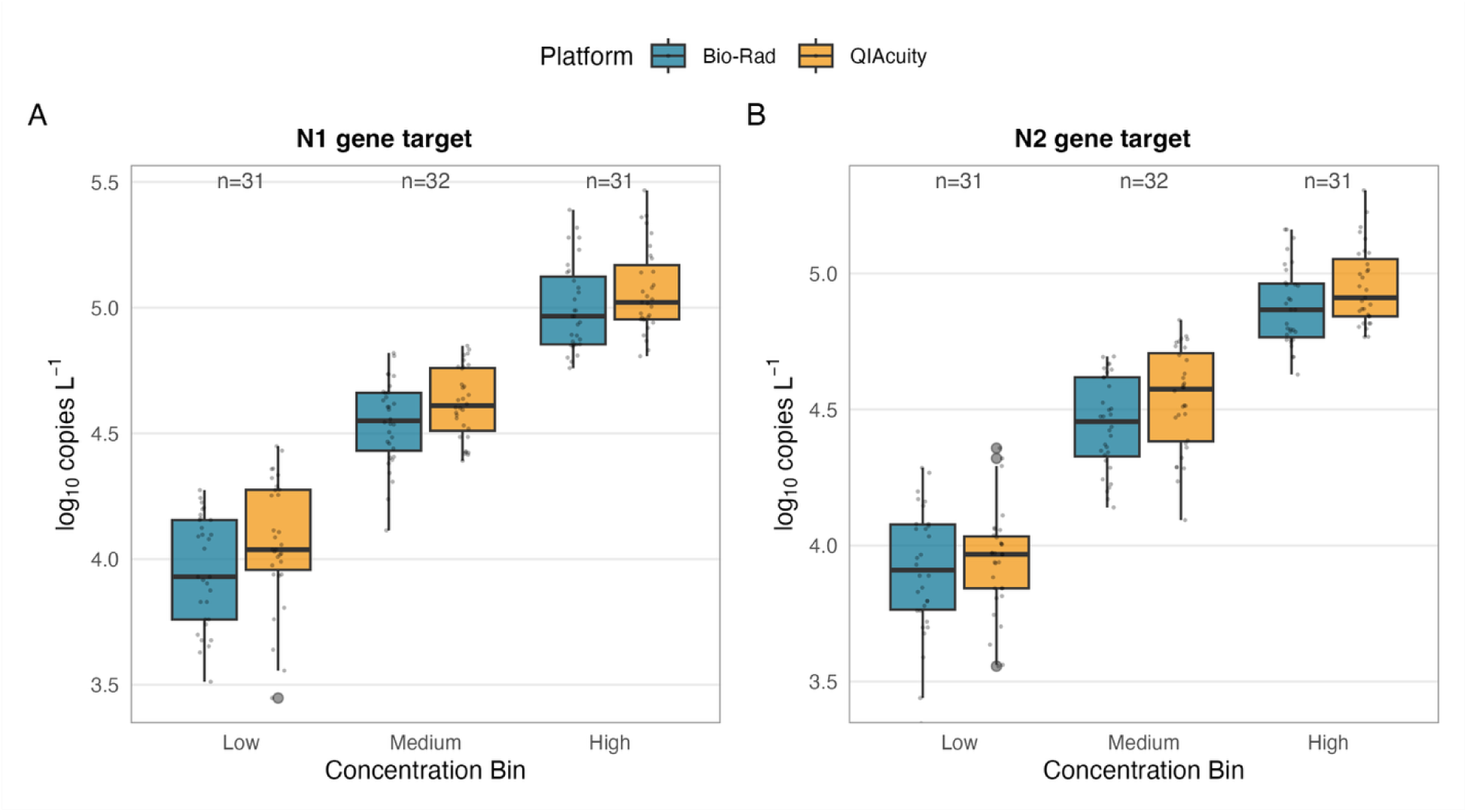
Platform comparison of SARS-CoV-2 quantification stratified by concentration bin. Box plots showing distribution of log_10_-transformed concentrations (copies L^-1^) for (A) N1 gene target and (B) N2 gene target across low, medium, and high concentration bins. Bio-Rad QX200 data shown in blue; QIAGEN QIAcuity data shown in green. Each box represents the interquartile range (IQR, 25^th^-75^th^ percentile), with the horizontal line indicating the medium. Whiskers extend to 1.5x IQR or the most extreme data point within this range. Individual data points are overlaid as semi-transparent dots to show data distribution (n=31 for high and low bins, n=32 for medium bin). Asterisks indicate statistically significant differences between platforms within each bin as determined by linear mixed effects modeling (*p < 0.05, **p < 0.01, ***p < 0.001). Note that all statistically significant differences are ≤ 0.13 log copies L^-1^. Sample numbers are indicated for each concentration bin. Both platforms successfully quantified 100% of samples across all bins.

### Precision and Reproducibility

Precision was assessed by calculating coefficients of variation (CoV) from triplicate hyperwelled reactions for each sample. Across the complete dataset, both platforms demonstrated comparable precision for all targets. For N1, mean CoV values were 4.21% (Bio-Rad) and 4.17% (QIAcuity); for N2, 4.17% (Bio-Rad) and 3.96% (QIAcuity); and for BCoV, 7.61% (Bio-Rad) and 7.48% (QIAcuity). Modified signed-likelihood ratio tests confirmed no significant differences in CoV equality between platforms for either N1 or N2 across any concentration bin (p > 0.05 for all comparisons, Table 2).

One notable exception occurred in the low concentration bin for N2, where CoV differed significantly between platforms (CoV = 5.71% for Bio-Rad vs. 4.84% for QIAcuity, p < 0.001). However, this difference represents only 0.87 percentage points and does not substantially affect data quality or interpretation at these concentrations. For all other target-bin combinations, CoV values remained within ± 1 percentage point between platforms.

The BCoV processing control exhibited the highest CoV values in the low concentration bin (11.26% for Bio-Rad, 11.18% for QIAcuity), but critically, both platforms showed nearly identical precision (p = 1.00), demonstrating consistent recovery across the entire analytical workflow regardless of platform choice.

Analysis of single-well (non-hyperwelled) reactions yielded similar conclusions, with no statistically significant differences in CoV between platforms for any target across any concentration bin (Table S12). This finding validates that platform equivalence holds whether samples are analyzed as individual replicates or merged for increased sensitivity.

An important observation emerged when examining CoV across individual wastewater treatment plant (WWTP) sites (Figure 3). Sites with high measurement variability showed consistently high CoV of both platforms, while sites with low variability maintained low CoV across both platforms (R^2^ = 0.89 for site-specific CoV correlation between platforms). This strong correlation indicates that variability is primarily driven by sample matrix characteristics or site-specific factors rather than platform-dependent analytical variability. The pattern held true across all three concentration bins, confirming that certain WWTPs consistently produce more heterogenous samples regardless of viral concentration or measurement platform.

**Figure 3.**
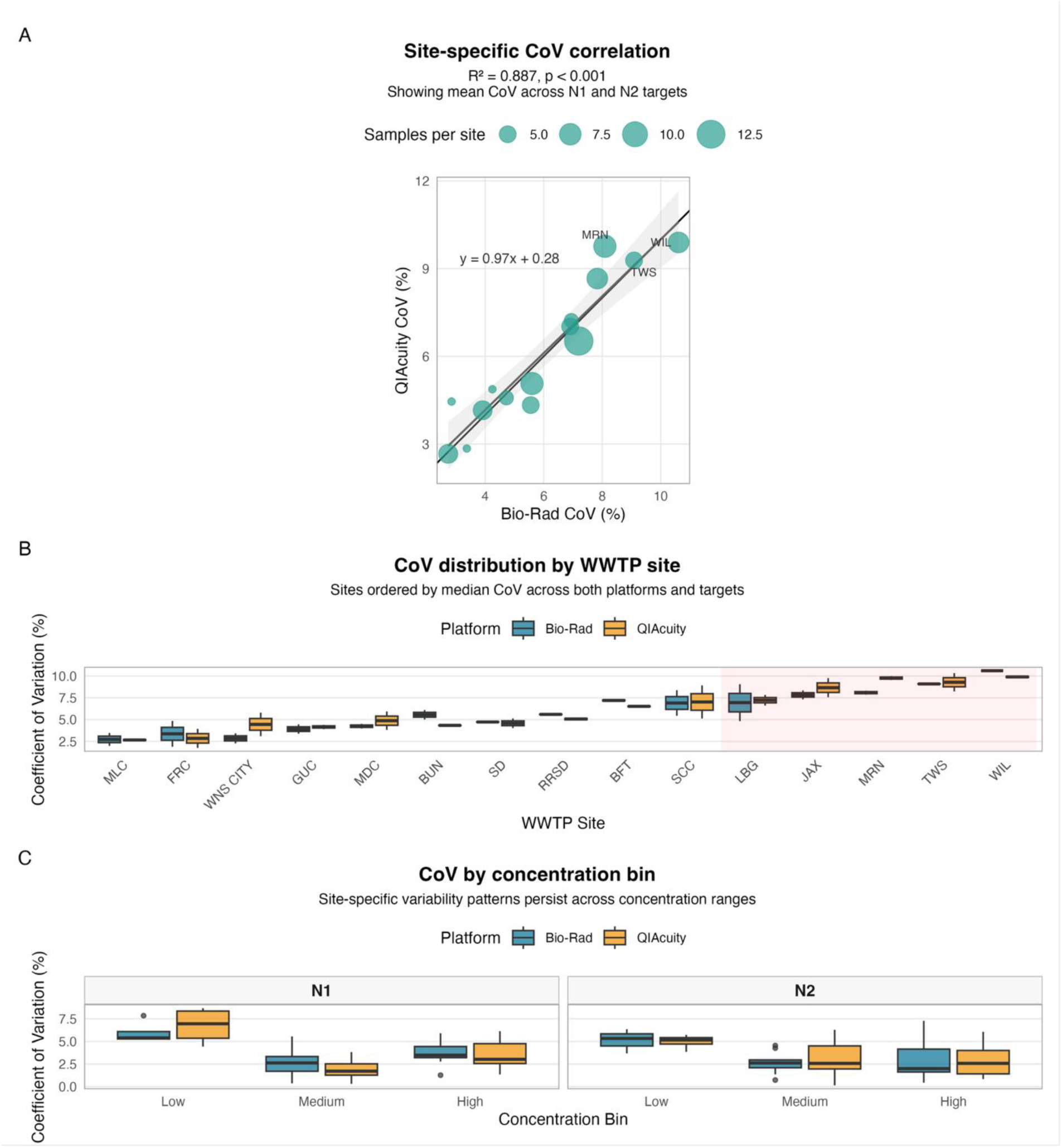
Site-specific coefficient of variation (CoV) demonstrates platform-independent measurement variability. (A) Scatterplot showing CoV (%) between Bio-Rad QX200 (x-axis) and QIAGEN QIAcuity (y-axis) platforms for individual wastewater treatment plant (WWTP) sites. Each point represents mean CoV across N1 and N2 targets for a single WWTP site (n=24 sites). Point size scaled by number of samples from each site. Solid black line represents 1:1 agreement; dashed line shows linear regression fit (R^2^ = 0.89, p<0.001). Sites with high variability on one platform show equivalently high variability on the other platform, indicating that measurement precision is driven by sample matrix characteristics rather than analytical platform. (B) Box plots showing CoV distribution by WWTP site for both platforms, with sites ordered by median CoV. Sites are color-coded consistently between platforms. Gray shading highlights the five sites with highest CoV, demonstrating consistent high variability across both platforms. (C) CoV values stratified by concentration bin (low/medium/high) for both platforms, showing that site-specific patterns persist across concentration ranges. Error bars represent standard error of the mean. No significant differences in CoV between platforms were observed for any site-bin combination (Modified signed-likelihood ratio test, p > 0.05 for all comparisons except N2 low bin).

### Limits of Detection

Following Clinical and Laboratory Standards Institute (CLSI) guidelines (document EP17-A2), we determined the limit of blank (LOB) using eight technical replicates of eight pre-pandemic wastewater samples (collected in 2018) that were confirmed negative for SARS-CoV-2. No positive partitions were detected in the N1 or N2 assays on either platform, establishing an LOB of zero for both platforms. This result confirms that both platforms produce no false-positive signals when properly optimized, providing confidence in low-level detections during routine surveillance.

Process limits of detections (PLOD) were determined through Probit analysis of 30 replicates across six serial dilutions, representing the concentration at which samples would be detected 95% of the time. The Bio-Rad QX200 achieved PLOD values of 2,160 copies L^-1^ for N1 and 2,680 copies L^-1^ for N2. The QIAcuity platform exhibited slightly higher PLOD values of 5,650 copies L^-1^ for N1 and 9,700 copies L^-1^ for N2 (Table 3).

**Table 3.** Limit of blank and process limit of detection for SARS-CoV-2 targets on both dPCR platforms.

| Platform | Target | LOB<br>(copies L<sup>-1</sup>)<sup>a</sup> | PLOD<br>(copies L<sup>-1</sup>)<sup>b</sup> | PLOD Fold<br>Difference<sup>c</sup> | Analytical LOD<br>(partitions)<sup>d</sup> |
| --- | --- | --- | --- | --- | --- |
| **Bio-Rad QX200** | N1 | 0 | 2,160 | -- | ≥3 positive droplets |
|  | N2 | 0 | 2,680 | -- | ≥3 positive droplets |
| **QIAGEN** | N1 | 0 | 5,650 | 2.6x | ≥3 positive partitions |
| **QIAcuity** | N2 | 0 | 9,700 | 3.6x | ≥3 positive partitions |

<sup>a</sup>Limit of blank (LOB) determined using eight technical replicates of eight pre-pandemic wastewater samples (collected 2018) confirmed negative for SARS-CoV-2. Zero positive partitions detected on both platforms for both targets, establishing LOB = 0 copies L<sup>-1</sup>.
<sup>b</sup>Process limit of detection (PLOD) determined by Probit analysis of 30 replicates across six serial dilutions following CLSI document EP17-A2 guidelines. PLOD represents concentration at which 95% detection probability is achieved across the entire workflow (filtration, extraction, RT, and dPCR).
<sup>c</sup>Fold difference calculated as QIAcuity PLOD / Bio-Rad PLOD.
<sup>d</sup>Analytical limit of detection applied consistently across both platforms after merging triplicate reactions. Samples with ≥3 total positive partitions across merged triplicates considered positive.
Both PLOD values fall below typical concentrations during community SARS-CoV-2 transmission (>10<sup>4</sup> copies L<sup>-1</sup>). Detection rate: 100% for both platforms in low concentration bin (1x10<sup>3</sup> to 5x10<sup>3</sup> copies L<sup>-1</sup>, n=31).

The approximately 2.5-3.6-fold difference in PLOD between platforms likely reflects the slightly larger reaction volume analyzed by the Bio-Rad platform. However, both platforms demonstrated PLOD values well below the typical concentrations encountered during community transmission of SARS-CoV-2 (generally >10^4^ copies L^-1^), and both successfully detected 100% of samples in our low concentration bin (1×10^3^ to 5×10^3^ copies L^-1^). The analytical limit of detection, defined as three positive partitions per well after merging triplicates, was applied consistently across both platforms and proved adequate for sensitive detection across all concentration ranges tested.

### Workflow and Cost Considerations

We conducted a comprehensive time-motion study comparing hands-on time, instrument time, and total time-to-result for processing 96 wells of wastewater samples on both platforms (Table 4). Hands-on time for mastermix preparation differed substantially: 20 minutes for the Bio-Rad platform (single 96-well plate) versus 80 minutes for the QIAcuity platform (four 24-well nanoplates required for equivalent throughput). While partition formation required zero hands-on time for both platforms, the Bio-Rad Automated Droplet Generator (AutoDG) requires 45 minutes of instrument time per 96 wells, whereas the QIAcuity nanoplates contains pre-formed partitions, eliminating this step entirely.

**Table 4.**
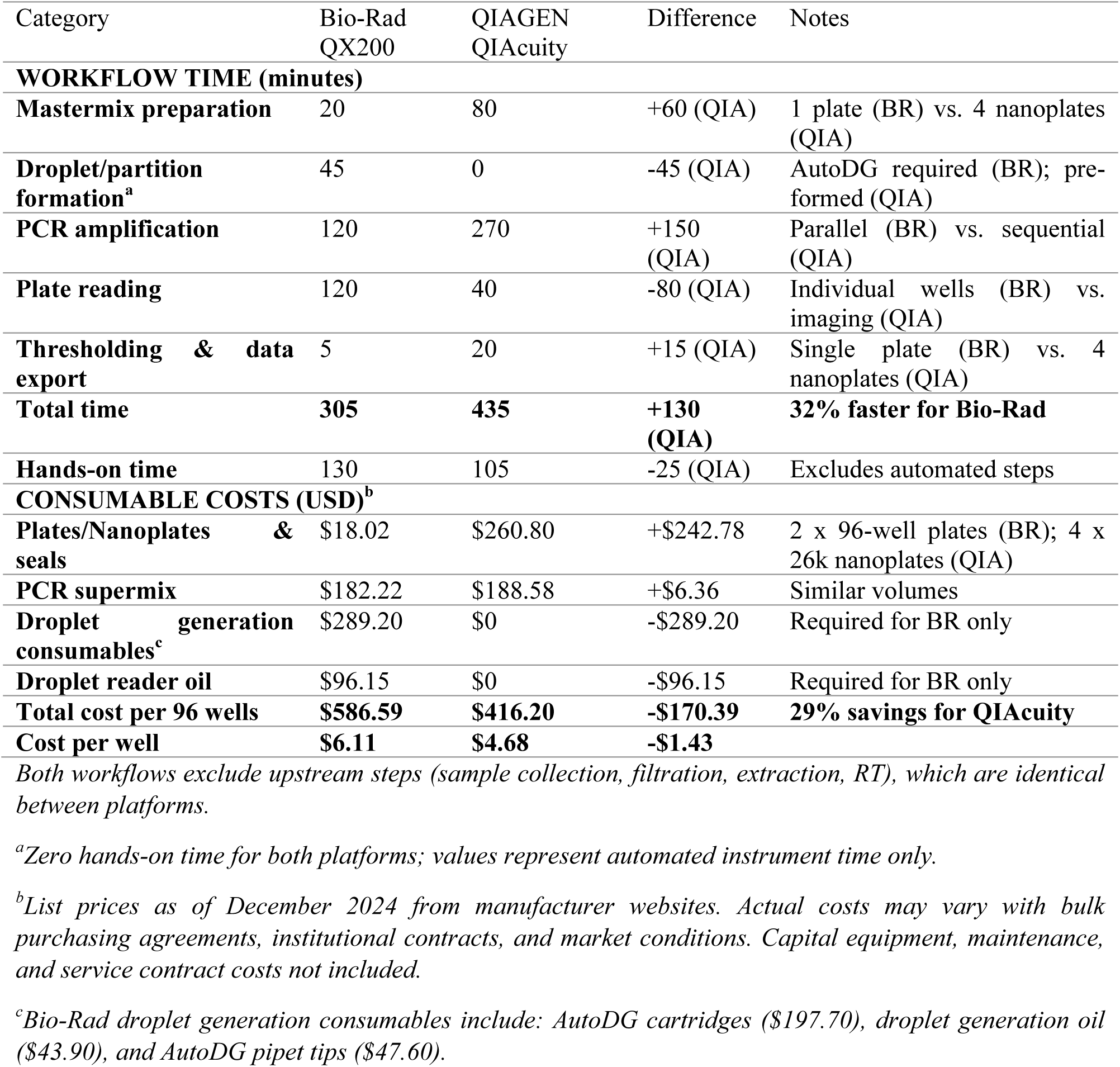
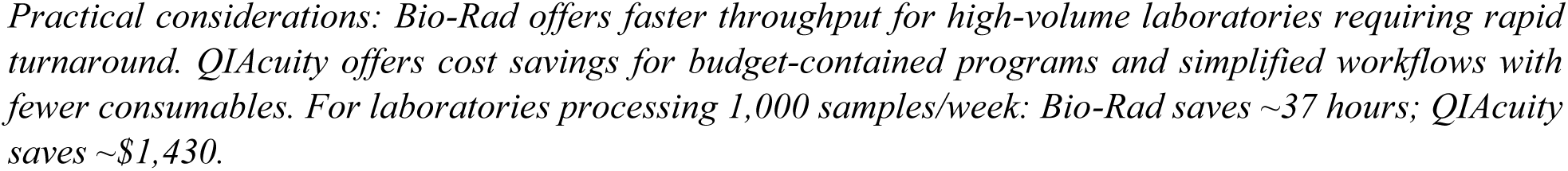
Workflow time and consumable cost comparison for processing 96 wells.

| Category | Bio-Rad QX200 | QIAGEN QIAcuity | Difference | Notes |
| --- | --- | --- | --- | --- |
| **WORKFLOW TIME (minutes)** |  |  |  |  |
| **Mastermix preparation** | 20 | 80 | +60 (QIA) | 1 plate (BR) vs. 4 nanoplates (QIA) |
| **Droplet/partition formation<sup>a</sup>** | 45 | 0 | -45 (QIA) | AutoDG required (BR); pre-formed (QIA) |
| **PCR amplification** | 120 | 270 | +150 (QIA) | Parallel (BR) vs. sequential (QIA) |
| **Plate reading** | 120 | 40 | -80 (QIA) | Individual wells (BR) vs. imaging (QIA) |
| **Thresholding & data export** | 5 | 20 | +15 (QIA) | Single plate (BR) vs. 4 nanoplates (QIA) |
| **Total time** | **305** | **435** | **+130 (QIA)** | **32% faster for Bio-Rad** |
| **Hands-on time** | 130 | 105 | -25 (QIA) | Excludes automated steps |
| **CONSUMABLE COSTS (USD)<sup>b</sup>** |  |  |  |  |
| **Plates/Nanoplates & seals** | \$18.02 | \$260.80 | +\$242.78 | 2 x 96-well plates (BR); 4 x 26k nanoplates (QIA) |
| **PCR supermix** | \$182.22 | \$188.58 | +\$6.36 | Similar volumes |
| **Droplet generation consumables<sup>c</sup>** | \$289.20 | \$0 | -\$289.20 | Required for BR only |
| **Droplet reader oil** | \$96.15 | \$0 | -\$96.15 | Required for BR only |
| **Total cost per 96 wells** | **\$586.59** | **\$416.20** | **-\$170.39** | **29% savings for QIAcuity** |
| **Cost per well** | **\$6.11** | **\$4.68** | **-\$1.43** | |

Both workflows exclude upstream steps (sample collection, filtration, extraction, RT), which are identical between platforms.
<sup>a</sup>Zero hands-on time for both platforms; values represent automated instrument time only.
<sup>b</sup>List prices as of December 2024 from manufacturer websites. Actual costs may vary with bulk purchasing agreements, institutional contracts, and market conditions. Capital equipment, maintenance, and service contract costs not included.
<sup>c</sup>Bio-Rad droplet generation consumables include: AutoDG cartridges (\$197.70), droplet generation oil (\$43.90), and AutoDG pipet tips (\$47.60).
*Practical considerations: Bio-Rad offers faster throughput for high-volume laboratories requiring rapid turnaround. QIAcuity offers cost savings for budget-contained programs and simplified workflows with fewer consumables. For laboratories processing 1,000 samples/week: Bio-Rad saves ~37 hours; QIAcuity saves ~\$1,430.*

PCR amplification times favored the QIAcuity platform for small batches but favored the Bio-Rad platform for full 96-well runs. A single 24-well QIAcuity nanoplate requires 90 minutes for thermal cycling, with subsequent plates processed sequentially at 60 minutes each, totaling approximately 270 minutes for 96 wells. In contrast, the Bio-Rad platform processes all 96 wells simultaneously in 120 minutes. The plate reading step showed the opposite pattern; the Bio-Rad QX200 reads individual wells at 1.25 minutes per well (120 minutes total for 96 wells), while the QIAcuity captures entire 24-well plates via imaging in approximately 10 minutes (40 minutes total for 96 wells).

Overall, processing 96 wells from mastermix preparation through data export required approximately 305 minutes on the Bio-Rad platform compared to 435 minutes on the QIAcuity platform, representing a 130-minute (32%) time savings for Bio-Rad when processing full 96-well batches (Table 4). However, for laboratories processing smaller batches (≤ 24 samples), the QIAcuity platform’s time advantages in imaging and elimination of droplet generation may offset the longer thermal cycling time.

Cost analysis focused on consumables required per 96-well equivalent of testing, excluding costs for sample collection, extraction, and reverse transcription, which are identical for both platforms (Table 4). The Bio-Rad platform requires seven consumable items: AutoDG cartridges, droplet generation oil, AutoDG pipet tips, 96-well plates, pierceable foil seals, droplet reader oil, and PCR supermix, totaling $586.59 per 96-well plate ($6.11 per well) at December 2024 list prices. The QIAcuity platform requires only two consumables: nanoplates with seals and PCR supermix, totaling $416.20 per 96-well equivalent ($4.68 per well). This represents a cost savings of $170.39 per 96 wells ($1.43 per well, approximately 29%) for the QIAcuity platform.

The cost difference primarily reflects the Bio-Rad platform’s requirements for consumables associated with droplet generation and individual well reading. However, cost considerations must be balanced against capital equipment expenses (not analyzed in this study), maintenance costs, and the specific throughput needs of individual laboratories. Additionally, consumable costs fluctuate with vendor negotiations, bulk purchasing agreements, and market conditions, and the values presented here represent list prices that may not reflect actual procurement costs for surveillance programs.

## Discussion

Since the beginning of the SARS CoV-2 pandemic, more than 100 papers have used quantitative PCR (qPCR) or digital PCR (dPCR) platforms to quantify CDC-recommended SARS CoV-2 targets in wastewater influent and solids. While numerous publications have compared qRT-PCR and qRT-ddPCR based quantification of SARS CoV-2 targets and controls (14,21,26), to our knowledge, no studies have directly compared two digital PCR platforms for wastewater surveillance. In recognition of the demonstrated advantages of dPCR - including improved sensitivity for respiratory viral targets, reduced susceptibility to inhibition relative to qPCR, and the ability to merge replicate reactions (“hyper-welling”) to enhance detection of low-abundance or emerging targets - the CDC has recommended the use of digital PCR and provides technical support for both the QIAGEN QIAcuity dPCR and the Bio-Rad ddPCR platforms within wastewater surveillance programs (28). Here, we present the first blinded, direct comparison of these two dPCR platforms using wastewater influent samples spanning a wide range of concentrations for SARS-CoV-2 N1, N2, and bovine coronavirus (BCoV) gene targets.

Across 95 wastewater influent samples collected throughout North Carolina during 2021-2022 and spanning three orders of magnitude in viral concentration, we observed no significant differences in either concentration estimates or measurement variability between the two platforms.

Mean differences between platforms were small (≤ 0.12 log copies L^-1^) for all targets, with strong linear correlations (R^2^ > 0.93) and regression slopes near unity. Coefficients of variation were comparable between platforms across all concentration ranges tested. These findings support the current practice in the CDC National Wastewater Surveillance System (NWSS) of accepting data from either platform interchangeably for public health decision-making. In addition, our evaluation of processing time and consumable costs showed that overall operational demands were broadly comparable between platforms, despite differences in specific workflow steps.

To date, we are aware of only one other study that has directly compared ddPCR and dPCR platforms for identical targets, focusing on cancer-associated DNA mutations in circulating free DNA from liquid biopsy samples (29). That study reported moderate platform-dependent differences, with dPCR demonstrating greater sensitivity for some low-abundance targets. These findings underscore that platform performance may vary depending on the molecular target, target abundance, analytical purpose, and sample matrix. Given the growing interest in expanding wastewater surveillance beyond SARS-CoV-2 to include additional viral, bacterial, and antimicrobial resistance targets, further direct comparisons of digital PCR platforms across a broader range of applications will be valuable.

A key finding of this study is that measurement variability was more strongly associated with wastewater treatment plant site than with analytical platform, highlighting the influence of sample matrix characteristics on quantitative precision. Wastewater is a highly heterogeneous matrix, and prior studies have demonstrated that differences in organic load, industrial input, and treatment processes can substantially affect analytical variability regardless of molecular platform (19,20,25). Sites exhibiting high coefficients of variation on one platform showed equivalently high variation on the other platform (R^2^ = 0.89), and this pattern persisted across all concentration bins. Identifying high-variability sites through initial characterization studies could inform sampling strategy modifications (e.g., composite sampling over longer time periods, increased sampling frequency, or collection at different locations within the treatment plant) to improve data quality. Our findings reinforce the conclusion that platform choice alone is unlikely to drive differences in reported concentrations and underscore the importance of standardized sampling, processing, and quality control practices across surveillance networks.

The demonstration of platform equivalence across low, medium, and high concentration bins is particularly significant for wastewater surveillance applications, where sample concentrations can vary dramatically based on community transmission levels, population served, and temporal dynamics. Both platforms successfully detected and quantified 100% of samples in the low concentration bin (1×10^3^ to 5×10^3^ copies L^-1^), with only minor differences in precision. While the Bio-Rad QX200 exhibited lower process limits of detection (2,160 – 2,680 copies L^-1^) compared to the QIAcuity (5,650 – 9,700 copies L^-1^), both values fall well below concentrations typically encountered during community transmission. The 2.5 – 3.6-fold difference in PLOD likely reflects differences in analyzed volume rather than fundamental differences in platform sensitivity. Importantly, neither platform produced false-positive signals in pre-pandemic samples, establishing confidence in detection specificity across both platforms.

The small but statistically significant differences observed between platforms in certain concentration bins (0.04 – 0.13 log copies L^-1^) warrant discussion. While statistical tests identified these differences, their magnitude is substantially smaller than the typical day-to-day variability observed in wastewater surveillance (often 0.3 – 0.5 log copies L^-1^) and far below the variation associated with biological and environmental factors affecting viral shedding and fate in sewage systems. Public health interpretation of wastewater data focuses on trend directionality and relative changes rather than absolute concentrations, given the numerous sources of uncertainty inherent in wastewater-based epidemiology. From this perspective, differences of 0.04 – 0.13 log units would not alter public health conclusions or trigger different response actions. The absence of platform x target interactions further reinforces that any observed differences are systematic and would not affect relative comparisons over time within a surveillance program using a single platform.

The methodological rigor of this study strengthens confidence in our findings. Blinded sample selection eliminated potential bias in sample choice or data interpretation. The use of archived samples from routine surveillance ensured analysis of authentic wastewater matrices across genuine concentration ranges encountered in operational programs. Comprehensive quality control, including processing controls (BCoV), extraction efficiency controls, reverse transcription controls, and PCR inhibition controls, verified that both platforms performed equivalently throughout the entire analytical workflow. Our approach to fluorescence thresholding, which used consistent criteria across platforms and accounted for N1 mutation-related population heterogeneity, provides a replicable framework for other laboratories conducting platform comparisons. The inclusion of both hyperwelled and single-well analyses confirmed that platform equivalence holds regardless of whether laboratories choose to merge technical replicates for improved sensitivity at low concentrations.

In addition to analytical performance, operational considerations such as processing time and consumable costs are important factors for laboratories engaged in sustained surveillance efforts. In this study, the Bio-Rad QX200 processed 96 samples approximately 32% faster (305 vs. 435 minutes), primarily due to simultaneous thermal cycling of all wells and the elimination of the sequential processing required by the QIAcuity’s 24-well format. However, the QIAcuity platform offered approximately 29% lower consumable costs ($4.68 vs. $6.11 per well), reflecting its simpler workflow with pre-formed partitions and imaging-based detection. These differences translate to meaningful impacts at scale; a laboratory processing 1,000 samples per week would save approximately 37 hours using the Bio-Rad platform or approximately $1,430 per week using the QIAcuity platform. These differences reflect distinctions in platform design rather than analytical limitations and may inform platform selection based on laboratory-specific capacity, sample throughput requirements, and budgetary constraints.

Several limitations of this study should be acknowledged. First, our comparison focused exclusively on filtered wastewater influent samples and well-characterized SARS-CoV-2 N1 and N2 targets. Since the completion of this study, many CDC NWSS laboratories have transitioned to alternative SARS-CoV-2 targets or expanded surveillance to include additional respiratory pathogens (influenza, RSV, human metapneumovirus) and other targets of public health interest. Platform performance may differ for other targets, particularly those with different abundance and inhibition profiles. Second, all samples originated from WWTPs participating in the North Carolina Wastewater Monitoring Network, which primarily serves municipal systems in a single geographic region. Performance in other matrices (e.g., primary settled solids, untreated septage, or samples from unique sewershed characteristics) may differ. Third, our time and cost analyses represent a snapshot of 2024 conditions and specific laboratory configurations (Auto-DG for Bio-Rad, four-nanoplate capacity for QIAcuity). Costs and workflows evolve with updated instrumentation, revised protocols, and changing market conditions. Finally, we did not evaluate inter-laboratory variability, an important consideration as wastewater surveillance expands to diverse laboratory settings with varying staff expertise and quality management systems.

As wastewater surveillance transitions from pandemic emergency response to routine public health infrastructure, establishing confidence in data comparability across platforms, laboratories, and jurisdictions becomes essential. Our findings demonstrate that the Bio-Rad QX200 and QIAGEN QIAcuity platforms produce equivalent quantitative data for SARS-CoV-2 surveillance in wastewater, supporting their interchangeable use in programs such as the CDC NWSS. This flexibility is essential for sustaining broad participation across jurisdictions and laboratory types as wastewater surveillance continues to evolve from an emergency response measure to a durable component of public health infrastructure.

## Acknowledgements

We gratefully acknowledge the North Carolina Department of Health and Human Services (NCDHHS) and Centers for Disease Control and Prevention (CDC) for their support of the NC WWMN. We extend our appreciation to the WWTP operators and staff across North Carolina, and all members of the Noble lab who collected and processed samples as part of routine surveillance efforts, making this study possible. We thank Dr. Sandra McLellan for providing pre-pandemic wastewater samples used in LOB determinations. We are grateful to Bio-Rad Laboratories and QIAGEN for providing technical consultation, application support, and reagents during the course of this study.

Initial funding was received by R.T. Noble for a Pilot COVID-19 Wastewater Surveillance Project from the North Carolina Policy Collaboratory. Additional follow-on support was received for the project entitled “Tracking SARS-CoV-2 in the Wastewater Across a Range of North Carolina Municipalities” which was also funded by the North Carolina Policy Collaboratory and provided important opportunities for assay optimization across both platforms. Additional support was provided by the North Carolina Department of Health and Human Services Division of Public Health for collaborative work with wastewater utilities across the State of North Carolina. We thank the work of the wastewater utilities across the State of North Carolina for their participation in the NC Wastewater Monitoring Network. The funders had no role in study design, data collection and analysis, decision to publish, or preparation of the manuscript.

## Data Availability

All data supporting the findings of this study are available within the manuscript and its Supplemental Materials.

